# Longitudinal serology in SARS-CoV-2 infected individuals in India – a prospective cohort study

**DOI:** 10.1101/2021.02.04.21251140

**Authors:** Ramachandran Thiruvengadam, Souvick Chattopadhyay, Farha Mehdi, Bapu Koundinya Desiraju, Susmita Chaudhuri, Savita Singh, Vandita Bhartia, Pallavi Kshetrapal, Uma Chandra Mouli Natchu, Nitya Wadhwa, Shailaja Sopory, Mudita Wahi, Anil K. Pandey, Asim Das, Nidhi Anand, Nandini Sharma, Pragya Sharma, Sonal Saxena, Deepa Sindhu, Brahmdeep Sindhu, Dharmendra Sharma, Navin Dang, Gaurav Batra, Gagandeep Kang, Shinjini Bhatnagar, for DBT India Consortium for COVID-19 Research

**Affiliations:** Translational Health Science and Technology Institute, Faridabad, Haryana, India; St. John’s Medical College and St. John’s Research Institute, Bengaluru, India; ESIC Medical College and Hospital, Faridabad, Haryana, India; Maulana Azad Medical College and Lok Nayak Hospital, New Delhi, India; Civil Hospital, Gurugram, India; Civil Hospital, Palwal, India; Dr Dang’s Laboratory, New Delhi, India; Christian Medical College, Vellore, India

**Keywords:** COVID-19, serology, severe COVID-19, asymptomatic, cohort

## Abstract

Clinical and epidemiological characteristics of SARS-CoV-2 infection are now widely available, but there are few data on longitudinal serology in large cohorts, particularly from low-and middle-income countries. We established an ongoing prospective cohort of 3840 SARS-CoV-2 RT-PCR positive individuals in the Delhi-National Capital Region of India, to document clinical and immunological characteristics during illness and convalescence. The IgG responses to the receptor binding domain (RBD) and nucleocapsid were assessed at 0-7, 10-28 days and 6-10 weeks after infection. The clinical predictors of seroconversion were identified by multivariable regression analysis. The seroconversion rates in the post-infection windows of 0–7 days, 10–28 days and 6–10 weeks were 46%, 84.7% and 85.3% respectively (n=782). The proportion with a serological response increased with severity of COVID-19 disease. All participants with severe disease, 89.6% with mild to moderate infection and 77.3% of asymptomatic participants had IgG antibodies to the RBD antigen. The threshold values in the nasopharyngeal viral RNA RT-PCR in a subset of asymptomatic and symptomatic seroconverters were comparable (p value: 0.48), with similar results among non-seroconverters (p value: 0.16) (n=169). This is the first report of longitudinal humoral immune responses to SARS-CoV-2 infection over a period of ten weeks from South Asia. The low seropositivity in asymptomatic participants and differences between assays highlight the importance of contextualizing the understanding of population serosurveys.

**Summary:** We measured anti-SARS-CoV-2 RBD and NC protein IgG in a multi-hospital-based prospective cohort from northern India up to ten weeks post-infection. The lower seroconversion rate among asymptomatic RT-PCR positive participants has public health significance particularly for interpreting community seroprevalence estimates.

## Introduction

Coronavirus disease 2019 (COVID-19), caused by SARS-CoV-2, has emerged as one of the most significant global public health challenges in the twenty-first century. The clinical phenotype of SARS-CoV-2 infection varies across a spectrum of asymptomatic to mild (moderate) illness to severe COVID-19. The proportion of the specific phenotypes differs with geographical location [1]. Prospectively followed cohorts of SARS-CoV-2 infected individuals provide an opportunity to accurately describe the clinical characteristics of the disease in diverse geographical settings over different time periods of evolution of the pandemic. Such cohorts when combined with a systematic collection of biospecimens can serve as useful platforms to answer emerging questions of public health importance.

The characterization of immunological response to SARS-CoV-2 infection and its association with the clinical spectrum of disease is still evolving. Detection of anti-SARS-CoV-2 antibodies targeting nucleocapsid (NC) and spike (S) protein, particularly the receptor binding domain (RBD) of the S protein is being used to evaluate serological humoral responses by age and clinical phenotypes. Data on persistence of antibodies against these proteins is emerging, but few studies are from low- and middle-income countries (LMICs).

We measured IgG antibodies against SARS-CoV-2 RBD and NC protein in participants enrolled into a multi-hospital based prospective cohort from northern India up to at least six weeks post-infection and correlated the clinical and demographic differences between seroconverters and non-seroconverters.

## Methods

### Study design and participants

This study was developed by an interdisciplinary of research institutes and hospitals in the National Capital Region (NCR), India; coordinated by the Translational Health Science and Technology Institute. The main clinical sites are ESIC Medical College Hospital, Faridabad and Loknayak Hospital, New Delhi. The study protocol was approved by the Institute Ethics Committees of all participating institutions.

### Cohort enrolment

In the ongoing cohort, we enrolled COVID-19 positive patients across all ages tested or admitted in designated COVID-19 testing centres or hospitals within five days of their positive RT-PCR test. The eligibility criteria was a positive RT-PCR done as per the National Testing Strategy of India and written informed consent [2]. The strategy to include participants from testing centres was to enrol outpatients while the in-patient recruitment in hospital wards was to enrich the cohort with patients with COVID-19 disease.

### Follow up

The follow-up visits were designed to capture the clinical outcomes of illness (10-28 days after being diagnosed with SARS-CoV-2 infection), early (6-10 weeks) and late (6 and 12 months) convalescent period. The duration of illness was defined as the date of onset of symptoms in symptomatic participants and from the date of testing positive for SARS-CoV-2 infection among those who were asymptomatic [3]. This report presents data up to early convalescence as later follow up is ongoing.

### Clinical data and biospecimen collection

A trained research team collected clinical data and biospecimens at enrolment and follow-up. The demography, clinical characteristics focussed on symptoms, comorbidities, drug history, and treatment details were collected by electronic data capture based on *a priori* developed SOP. Venous blood samples were collected, and transported following biosafety protocols recommended by the Government of India and the World Health Organization [4]. Serum was separated and stored in the biorepository at THSTI for subsequent analyses.

### Serological assays

#### Anti-RBD IgG ELISA

Anti-SARS-CoV-2 RBD IgG antibody was detected using an ELISA method as described earlier [5]. Briefly, samples and controls were diluted to 1:50 in the sample diluent, and 100μl was added per well of stabilized RBD coated plate. After 30 min of incubation at room temperature (RT) (23±2°C), the plates were washed six times with wash buffer. Fifty μl of ready to use conjugate (HRP-labelled Goat anti-Human IgG Fcγ specific antibody) was added per well and incubated for 30 min at RT, followed by six washes. Hundred μl of TMB substrate was added to each well, incubated at RT for 10 minutes and the reaction was stopped with 100 μl per well of stop solution. The absorbance was measured on a microplate reader at 450 nm with 650 nm as a reference wavelength. The pooled negative control and pooled positive control were run on each plate. The cut-off from each plate was calculated by taking the average OD of triplicate of negative control and the addition of 0.2 to this value. Signal to the cut-off ratio (S/Co) was calculated as the ratio of OD value from the test sample to the cut-off value. S/Co ratio of ≥ 1 was considered positive.

#### Anti-NC IgG Chemiluminescence assay

The anti-NC IgG chemiluminescence assay (SARS-CoV-2 IgG) by Abbott Diagnostics Division, Ireland, was performed as per manufacturer’s instructions, with calibrator and positive and negative controls run before each batch of antibody testing as per manufacturer’s protocol. The results are reported by dividing the chemiluminescent signal from sample by the mean of chemiluminescent signals from three calibrator replicates. The default result unit is Index (S/C) which above a cut-off of 1.4 is considered positive.

### Statistical analysis

Seroconverters were defined as those who tested seropositive by either of the assay at least once during the study period. The clinical phenotypes of COVID-19 were stratified into severe, mild to moderate and asymptomatic based on their most severe symptoms and/or the need for treatment at enrolment or at any time in their follow up period. Participants were considered to have severe COVID-19 if they died or required either of the following, oxygen supplementation or ventilatory support, or treatment in the intensive care unit for cardiopulmonary or multiorgan dysfunction. Mild to moderate disease was defined by the presence of symptoms of COVID-19 but not fulfilling the criteria for severe disease. Those who did not report any symptoms at enrolment or throughout the follow-up period were classified as asymptomatic COVID-19. The clinical and epidemiological characteristics of the participants were described as median and interquartile range (IQR) for continuous and as percentages for categorical variables. The univariate analyses were performed using chi-squared test for comparison of proportions and Mann Whitney U test for difference in distributions between different clinical groups (seroconverters and non-seroconverters). Multivariable regression analysis was done to identify independent predictors of seroconversion. The difference in cycle threshold values between asymptomatic and symptomatic individuals in both seroconversion and non-seroconversion categories were tested for significance using Mann Whitney U test. All statistical analyses were performed using R.

## Results

### Baseline and clinical characteristics of the participants

The cohort has 3790 COVID-19 participants on December 18, 2020, with the majority of enrolled from Lok Nayak Hospital, New Delhi (45%) and ESIC Medical College Hospital, Faridabad (44.6%). We present clinical results of the first 2504 (2139 from the hospital and 365 from testing centres) participants enrolled between April 2020 to October 2020, who were 6-10 weeks post-diagnosis. The median age was 44 years (IQR: 30, 57) with more than two-thirds males. Nearly two-thirds presented with symptoms, most commonly fever (61%), cough (48%), breathlessness (35%), sore throat (27%) and bodyache (22%). Only one in five individuals had a history of primary contact defined as direct contact for 15 minutes without a mask with someone who tested COVID-19 positive. A history of secondary contact, defined as direct contact for 15 minutes without mask with someone who in turn had a primary contact history was noted in 2%. Among those who gave a contact history, the median duration since contact was four days (IQR: 2, 7). The baseline characteristics of the participants included for serological analysis (n=782) are provided in table -1.

**Table 1.** Demographic and clinical characteristics of participants.

| **Baseline Characteristics** | **Full cohort (N = 2504)** |  | **Participants included for serological evaluation (N= 782)** |
| --- | --- | --- | --- |
|  | **N** | **Median (IQR) or n(%)** |  |
| **Age in years**<br>**median (IQR)** | 2452 | 42 (29, 55) | 40 (29, 53) |
| **Male n%** | 2,504 | 1703 (68%) | 554 (71%) |
| **Symptomatic n%** | 2,504 | 2078 (83%) | 672 (86%) |
| **H/o primary contact n%** | 2,504 |  |  |
| No |  | 1510 (60%) | 449 (57%) |
| Not Known |  | 483 (19%) | 148 (19%) |
| Yes |  | 511 (20%) | 185 (24%) |
| **H/o secondary contact n%** | 2,504 |  |  |
| No |  | 1551 (62%) | 460 (59%) |
| Not Known |  | 889 (35%) | 298 (38%) |
| Yes |  | 64 (2.6%) | 24 (3.1%) |
| **Duration of contact in days**<br>**median (IQR)** | 386 | 4.0 (2.0, 7.8) | 4.0 (2.0, 8.0) |
| **Duration of contact in hours**<br>**per day**<br>**median (IQR)** | 309 | 12 (6, 24) | 12 (6, 24) |
| **Outcome characteristics** |  |  |  |
| **Disease severity<sup>1</sup> n%**<br>**(95%CI)** | 2,504 |  |  |
| Asymptomatic |  | 411 (16% 14.98,<br>17.92) | 107 (14% 11.35,<br>16.30) |
| Mild / Moderate |  | 1,638 (65%<br>63.51, 67.27) | 539 (69% 65.55,<br>72.15) |
| Severe |  | 455 (18% 16.67,<br>19.73) | 136 (17% 14.80,<br>20.23) |
| **Intensive care** | 2,278 | 75 (3.3% 2.59, 4.10) | 24 (3.1% 1.98, 4.53) |
| **Oxygen therapy** | 2,313 | 432 (19% 17.10, 20.32) | 134 (17% 14.55, 19.96) |
| **Mechanical ventilation** | 2,275 | 10 (0.4% 0.21, 0.80) | 2 (0.3% 0.03, 0.92) |
| **Death** | 2,504 | 23 (0.9% 0.58, 1.37) | 0 (0%) |
| **Time to death from date of diagnosis (days)** | 23 | 25 (16, 35) | NA |
| **Time to death from date of onset of symptoms (days)** | 20 | 34 (20, 43) | NA |

<sup>1</sup>Participants were considered to have severe COVID-19 if they died or required either of the following, oxygen supplementation or ventilatory support, or treatment in the intensive care unit for cardiopulmonary or multiorgan dysfunction. Mild to moderate disease was defined by the presence of symptoms of COVID-19 but not fulfilling the criteria for severe disease. Those who did not report any symptoms at enrolment or throughout the follow-up period were classified as asymptomatic COVID-19.

### Serological characteristics

The longitudinal serological evaluation included participants who contributed all three samples (n=782). The seroconversion rates in the windows 0–7 days, 10–28 days and 6–10 weeks were 46%, 84.7% and 85.3% respectively (Table 2). The proportion positive increased with severity of COVID-19 disease, 100% with severe disease, 89.6% with mild to moderate disease and 77.3% of asymptomatic positives had IgG antibodies to RBD antigen. As shown in Figure 1 and Table 3, the response to NC antigen by anti-NC CLIA was lower (severe–98.3%, mild–moderate 85.9%, and asymptomatic–69.9%). The higher seroconversion with disease severity was consistent when we evaluated the signal to cut-off ratio of both anti-RBD and anti-NC IgG antibody (Figure 2). Notably, those who were seropositive for anti RBD IgG at enrolment had almost 60% higher risk of severity (38/242 vs 29/294; RR: 1.59 (95% CI: 1.01, 2.50) than those who remained seronegative or became seropositive at a later time point. The threshold values in the nasopharyngeal viral RNA RT-PCR of asymptomatic and symptomatic seroconverters were comparable (p value: 0.48), as were asymptomatic and symptomatic non-seroconverters (p value: 0.16) (n=169) (Supplementary figure 2). In all those who mounted an IgG response, there was no demonstrable fall as seen by the signal-cut off ratio up to 10 weeks of follow up.

**Table 2.** Seropositivity for antibodies against different antigens (N=672)

|  | **Positive only**<br>**for anti-RBD**<br>**IgG** | **Positive only**<br>**for anti-NC**<br>**IgG** | **Positive for**<br>**both**<br>**antibodies** | **Positive for**<br>**either of the**<br>**antibodies** |
| --- | --- | --- | --- | --- |
| **Within 7 days of**<br>**illness** | 76 (11.3%) | 31 (4.6%) | 202 (30.1%) | 309 (46.0%) |
| **By 10-28 days of**<br>**illness** | 56 (8.3%) | 10 (1.5%) | 503 (74.9%) | 569 (84.7%) |
| **By 6-10 weeks of**<br>**illness** | 45 (6.7%) | 4 (0.6%) | 523 (77.8%) | 573 (85.3%) |

The seropositivity is represented as absolute numbers and percentages

**Table 3.** Proportion of seroconverters among different categories of disease severity.

| **Type of assay** | **Asymptomatic**<br>**n;%** | **Mild / Moderate**<br>**n;%** | **Severe n;%** | **Proportion among all categories n;%** |
| --- | --- | --- | --- | --- |
| Anti-RBD IgG<br>(N=743) | 160; 77.29%<br>(95%CI: 70.98, 82.81) | 420; 89.55%<br>(95%CI: 86.42, 92.17) | 67; 100.00%<br>(95%CI: 94.64, 100.00) | 647; 87%<br>(95%CI: 84.45, 89.41) |
| Anti-NC IgG<br>(N=673) | 130; 69.89%<br>(95%CI: 62.75, 76.39) | 364; 85.85%<br>(95%CI: 82.16, 89.02 (364) | 59; 98.33%<br>(95%CI: 91.06, 99.96) | 553; 82.16%;<br>(95%CI: 79.06, 84.99) |

The last column denotes the overall seropositivity rates.
The number in the paranthesis denotes the 95% confidence intervals of the percentage estimates od seroconverters.

**Figure 1.**
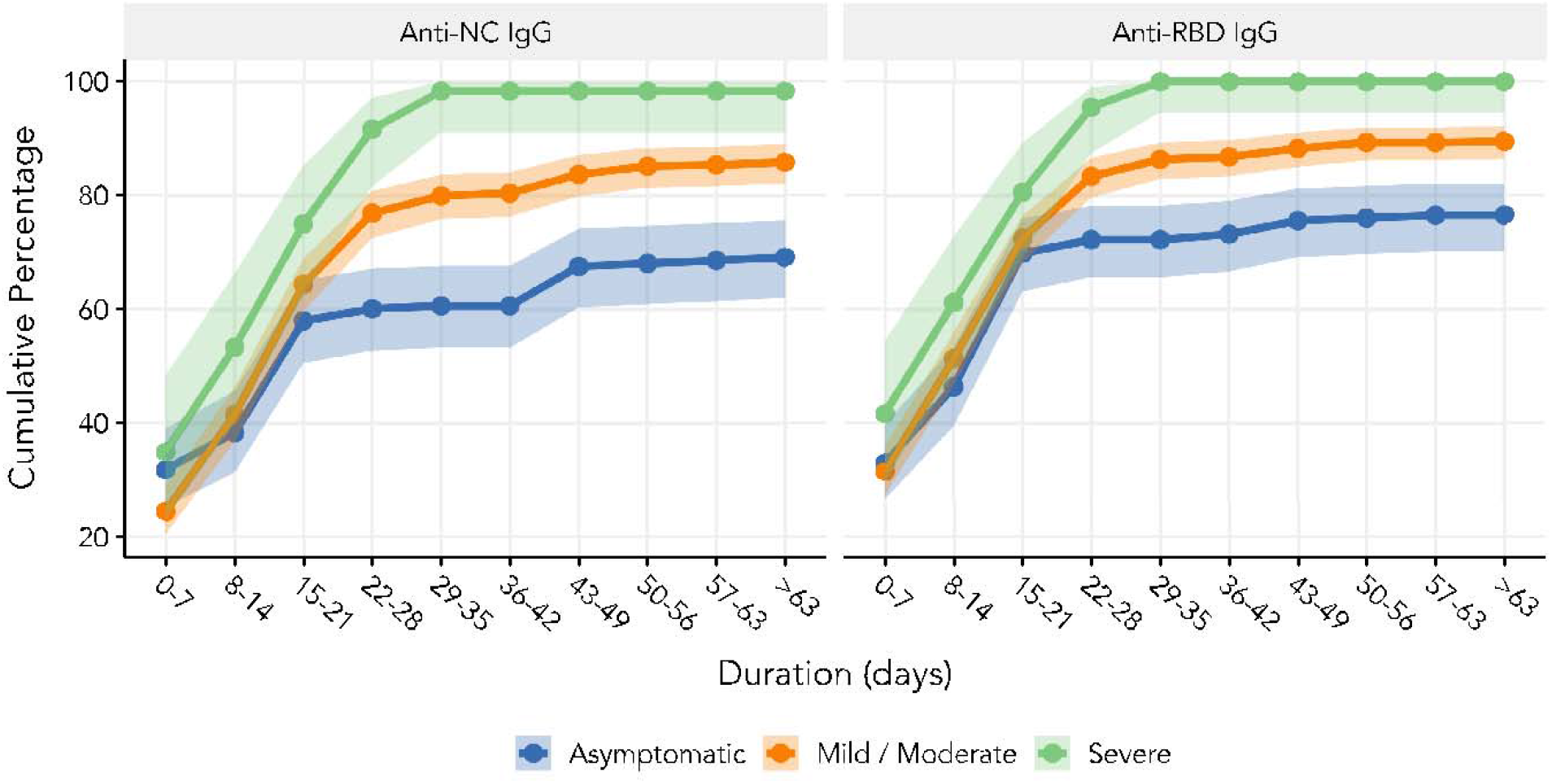
Cumulative seropositivity for anti-NC and anti-RBD IgG among different categories of disease severity. Anti-NC and anti-RBD IgG denote anti-nucleocapsid and anti-RBD Immunoglobulin-G respectively. The error bars indicate 95%CI of signal/cutoff ratio during that window of followup.

**Figure 2.**
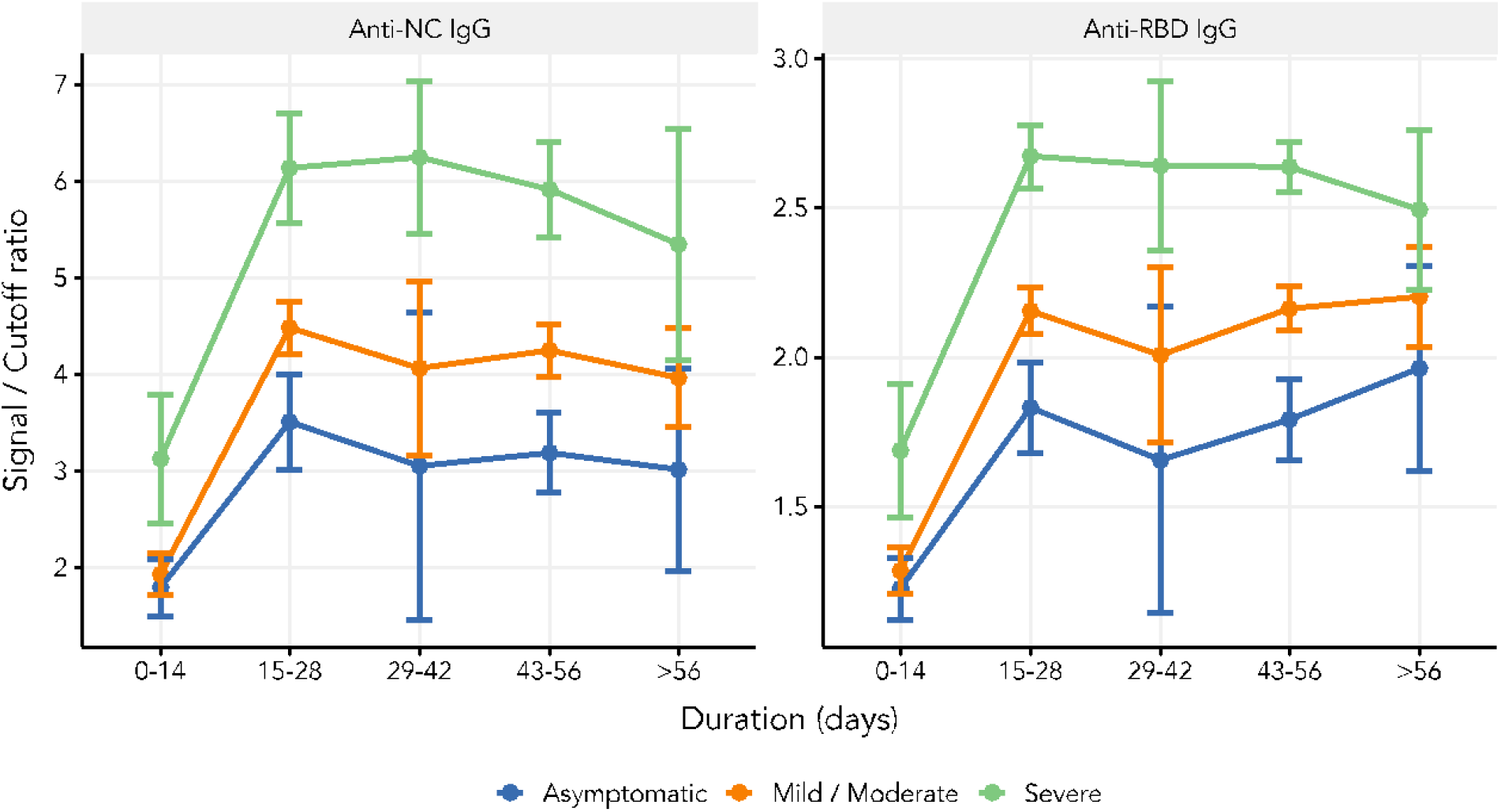
Longitudinal change in the signal to cut-off ratio among different categories of disease severity. Anti-NC and anti-RBD IgG denote anti-nucleocapsid and anti-RBD Immunoglobulin-G respectively. The error bars indicate 95% confidence interval of signal/cutoff ratio during that window of follow up.

### Predictors of seroconversion

Demographic and clinical characteristics associated with seroconversion are described in Table 4. More than 95% (n=105) participants over 60 years of age were IgG antibody positive; this proportion reduced with decreasing age. Interestingly, participants who reported no primary contact or those who were uncertain about the contact history seroconverted more than those who had a history of exposure. Seroconverters reported a longer exposure duration per day than those who did not show IgG response (median exposure: 24 hours vs 8 hours) (Table 5). Presence of pre-existing comorbidities such as chronic hypertension (p-value <0.001) and diabetes mellitus (p-value <0.001) were associated with seroconversion (Table 4). The multivariable model after adjusting for confounders showed that age (years) (aOR: 1.03; 95%CI: 1.02,1.05) and presence of symptoms at presentation (aOR: 3.23; 95%CI: 2.01,5.17) were independent predictors of seroconversion.

**Table 4:** Demographic and clinical characteristics associated with seroconversion.

| **Characteristic** | **N<sup>1</sup>** | **Proportion of**<br>**seroconverters**<br>**n(%)** | **p-value** |
| --- | --- | --- | --- |
| **Sex** |  |  |  |
| Female | 212 | 176 (83.02%) | 0.24 |
| Male | 513 | 445 (86.74%) |  |
| **Symptom status at enrolment** |  |  |  |
| Asymptomatic | 122 | 83 (68.03%) | <0.001 |
| Symptomatic | 603 | 538 (89.22%) |  |
| **Age** |  |  |  |
| 0-30 years | 190 | 143 (75.26%) | <0.001 |
| 30-60 years | 410 | 358 (87.32%) |  |
| > 60 years | 109 | 105 (96.33%) |  |
| **Comorbidities** |  |  |  |
| Asthma |  |  |  |
| No | 708 | 609 (86.02%) | 0.15 |
| Yes | 17 | 12 (70.59%) |  |
| Autoimmune disorders |  |  |  |
| No | 721 | 618 (85.71%) | 1.00 |
| Yes | 4 | 3 (75%) |  |
| Cancer |  |  |  |
| No | 715 | 612 (85.59%) |  |
| Yes | 10 | 9 (90%) |  |
| Diabetes |  |  |  |
| No | 597 | 500 (83.75%) | <0.001 |
| Yes | 128 | 121 (94.53%) |  |
| Heart Disease |  |  |  |
| No | 696 | 594 (85.34%) | 0.37 |
| Yes | 29 | 27 (93.1%) |  |
| Hypertension |  |  |  |
| No | 605 | 505 (83.47%) | <0.001 |
| Yes | 120 | 116 (96.67%) |  |
| Kidney disease |  |  |  |
| No | 717 | 616 (85.91%) | 0.17 |
| Yes | 8 | 5 (62.5%) |  |
| Liver Disease |  |  |  |
| No | 717 | 615 (85.77%) | 0.72 |
| Yes | 8 | 6 (75%) |  |
| More than one comorbidity |  |  |  |
| No | 624 | 530 (84.94 %) | 0.22 |
| Yes | 101 | 91 (90.1 %) |  |
| Smoking |  |  |  |
| No | 667 | 574 (86.06 %) | 0.39 |
| Yes | 58 | 47 (81.03 %) |  |
| Thyroid disorders |  |  |  |
| No | 697 | 597 (85.65 %) | 1.00 |
| Yes | 28 | 24 (85.71 %) |  |

<sup>1</sup>denotes the number of participants in the categories specified in column 1
The multivariable model including all the predictors listed in the table showed that age (years) (aOR: 1.03; 95%CI: 1.02,1.05) and presence of symptoms at presentation (aOR: 3.23; 95%CI: 2.01,5.17) were independent predictors of seroconversion.

**Table 5:** Differences in the epidemiological characteristics between seroconverters and non-seroconverters.

| Baseline Characteristics | N | Non seroconverters<sup>1</sup> | Seroconverters<sup>1</sup> | p-value |
| --- | --- | --- | --- | --- |
| Duration of contact (days) | 124 | 4.0 (1.0, 8.5)<br>(n = 31) | 4.0 (2.0, 7.0)<br>(n = 93) | 0.8 |
| Duration of contact in hours per day | 99 | 8 (2, 24)<br>(n = 27) | 24 (7, 24)<br>(n = 72) | 0.01 |
| Duration between date of first contact and date of diagnosis | 127 | 9 (5, 11)<br>(n = 33) | 6 (3, 10)<br>(n = 94) | 0.13 |
| Duration between date of last contact and date of diagnosis | 122 | 1.0 (-1.0, 6.0)<br>(n = 31) | 1.0 (-1.0, 4.0)<br>(n = 91) | 0.6 |

<sup>1</sup>Statistics presented: median (IQR)

## Discussion

Age and symptomatic status were independent positive predictors of seroconversion in our Indian cohort. The higher antibody response in severe COVID infection was expected. A severe infection indicates either a higher infectious dose [6] or spread of viruses beyond the respiratory tract. In both situations the probability of the host immune response increases [7–9]. Further, a longer exposure of the host immune system due to prolonged severe COVID infection may lead to generation of polyfunctional T cells, that elicit a cytokine storm and an evident humoral response[10-11]. Our findings differ from western reports that showed much higher seroconversion among asymptomatic individuals [12–14]. However, a recent study from Bangladesh has reported lower seroconversion rates (45%) among asymptomatic individuals [15]. This raises the possibility that the biological responses to SARS COV-2 may vary between different populations.

Age is a known strong predictor of severe COVID-19 and it is possible that in elderly individuals, there is systemic spread of infection, possibly due to an impaired innate immune protection. The presence of comorbidities such as chronic hypertension and diabetes mellitus were found to be associated with seroconversion in unadjusted analysis; a relationship probably confounded by age of the participant.

Nearly 15% of participants did not have IgG against both RBD and NC antigens. While the trends of seropositivity were largely similar, anti-RBD IgG were detectable marginally earlier in the course of illness and in a larger proportion of individuals than anti-NC IgG. This was particularly marked among the asymptomatic individuals. There could be many reasons for a poorer antibody response amongst asymptomatic individuals in our cohort. Unlike in severe infection, a possible lack of systemic spread of the virus in asymptomatic individuals would result in less exposure of the immune system. A large proportion of these individuals were identified by active contact tracing, and no or low seropositivity could reflect less exposure to a SARS-CoV-2 positive contact [16]. It has been shown that the nasopharyngeal viral load is significantly higher among individuals with severe COVID-19 as compared to mild illness [17].

The lower seroconversion rate among asymptomatic RT-PCR positive participants has critical public health significance particularly for interpreting community seroprevalence in highly affected geographical areas. Results from seroprevalence studies from India and other South Asian countries [18–22] may need to be adjusted in the light of our findings.

The signal-cut off ratio for both the assays reached their maxima between 15-28 days and plateaued suggesting a stable IgG response up to 10 weeks of illness. The longevity of IgG response has been reported to vary between 5 weeks to 4 months. [13, 22–25]. As shown in another global study [27], asymptomatic participants in our study did not show any decline of IgG to either antigens over a period of 10 weeks. This is in contrast to initial evidence showing early decline of IgG in asymptomatic infected individuals [3,13]. We will follow up the participants in this cohort for at least a year to evaluate the longevity of IgG response.

The major strengths of the study are the prospective data collection with high follow-up rates, and the use of two validated antibody assays. However, there are some limitations. Due to lack of robust IgM and IgA assays, we were unable to evaluate the immune response with respect to these isotypes. Further, due to the heterogeneity of RT-PCR assays used by the participating hospitals for molecular diagnosis, we do not have a comparable viral load data for all participants in our study. However, analysis in a subset of the individuals who had a similar RT-PCR assay showed no difference in nasopharyngeal viral load between seroconverters and non-seroconverters.

In conclusion, we systematically report clinical and epidemiological characteristics, and longitudinal humoral immune response to SARS-CoV-2 infection in a large Indian cohort. We will continue to study the kinetics of the immune response in a unique platform to evaluate the more complex and emerging questions on cellular immunity, reinfections, long COVID syndrome and population-level post immunization surveillance, when the cohort participants are immunized.

## Data Availability

The data referred to in the manuscript will be available on request after peer-reviewed publication, upon request to the corresponding author

## Author contributions

SB, GK, RT, UCM and GB conceptualized the study; All members of DBT India Consortium for COVID-19 Research devised methods, collected samples and clinical data; NW, UCM, RT, VB, MW, AKP, AD, NA, NS, PS, SSi, DSi and BS coordinated clinical site activities; GB, PK, SSo, SSi, SC, ND, SChat and FM provided reagents and analysed samples; RT, BKD and DSh curated and analysed data; SB, RT, GB, VB, SC, SSi, and PK drafted the manuscript; GK and SB coordinated and supervised the study. All authors contributed in revision and approved the final draft of the manuscript.

## Acknowledgments

We deeply thank the Department of Biotechnology, Government of India for supporting the consortium. We are grateful to the leadership and administration of all partner institutions in the consortium for their help and support. We thank all the clinical, laboratory and data management staff for their contributions to this work and the consortium at large.

## Members of DBT India consortium for COVID-19 Research

Coordinating Institute: Translational Health Science and Technology Institute (THSTI)

Coordinating Principal Investigator: Dr Shinjini Bhatnagar

Co- Principal Investigator: Dr Gagandeep Kang

Co-Investigators (Clinical): Drs Nitya Wadhwa, Uma Chandramouli Natchu, Ramachandran Thiruvengadam, Shailaja Sopory, Pallavi Kshetrapal, Bapu Koundinya Desiraju, Vandita Bhartia, Mudita Gosain, Monika Bahl

Co-Investigators (Laboratory): Drs Gaurav Batra, Guruprasad Medigeshi, Susmita Chaudhuri, Niraj Kumar, Tarun Sharma, Chandresh Sharma, Shailendra Mani, Tripti Shrivastava

International Centre for Genetic Engineering and Biotechnology (ICGEB): Dr Anmol Chandele

National Institute of immunology (NII): Drs. Amulya Panda, Nimesh Gupta

Maulana Azad Medical College and Lok Nayak Hospital, New Delhi: Drs. Nandini Sharma, Pragya Sharma, Sonal Saxena, J.C. Passey, Suresh Kumar

ESIC Medical College and Hospital, Faridabad, Haryana: Dr Anil K Pandey, Asim Das, Nikhil Verma, Needhi Anand, Sujata Roy Choudhary

Civil Hospital Gurugram (GCH), Haryana: Drs Deepa Sindhu, Jai Singh Malik

Civil Hospital Palwal (PCH), Haryana: Dr Brahmdeep Sindhu

Al-Falah School of Medical Science & Research Centre and Hospital, Dhouj, Haryana: Drs Bhupinder Kaur Anand, Shubham Girdhar

Medanta Hospital, Gurugram, Haryana: Drs Sushila Kataria, Pooja Sharma

Shaheed Hasan Khan Mewati Government Medical College, Nalhar, Nuh, Haryana: Dr Yamini

Lady Hardinge Medical College, New Delhi: Drs. Harish K. Pemde, Tanmaya Talukdar

SGT Medical college, Gurugram, Haryana: Drs. Pankaj Abrol, Mukesh Sharma

Dr. Dang’s Lab, New Delhi: Drs Navin Dang, Manavi Dang, Arjun Dang, Leena Chatterjee, Devjani De

## Supplementary information

**Supplementary figure 1.**
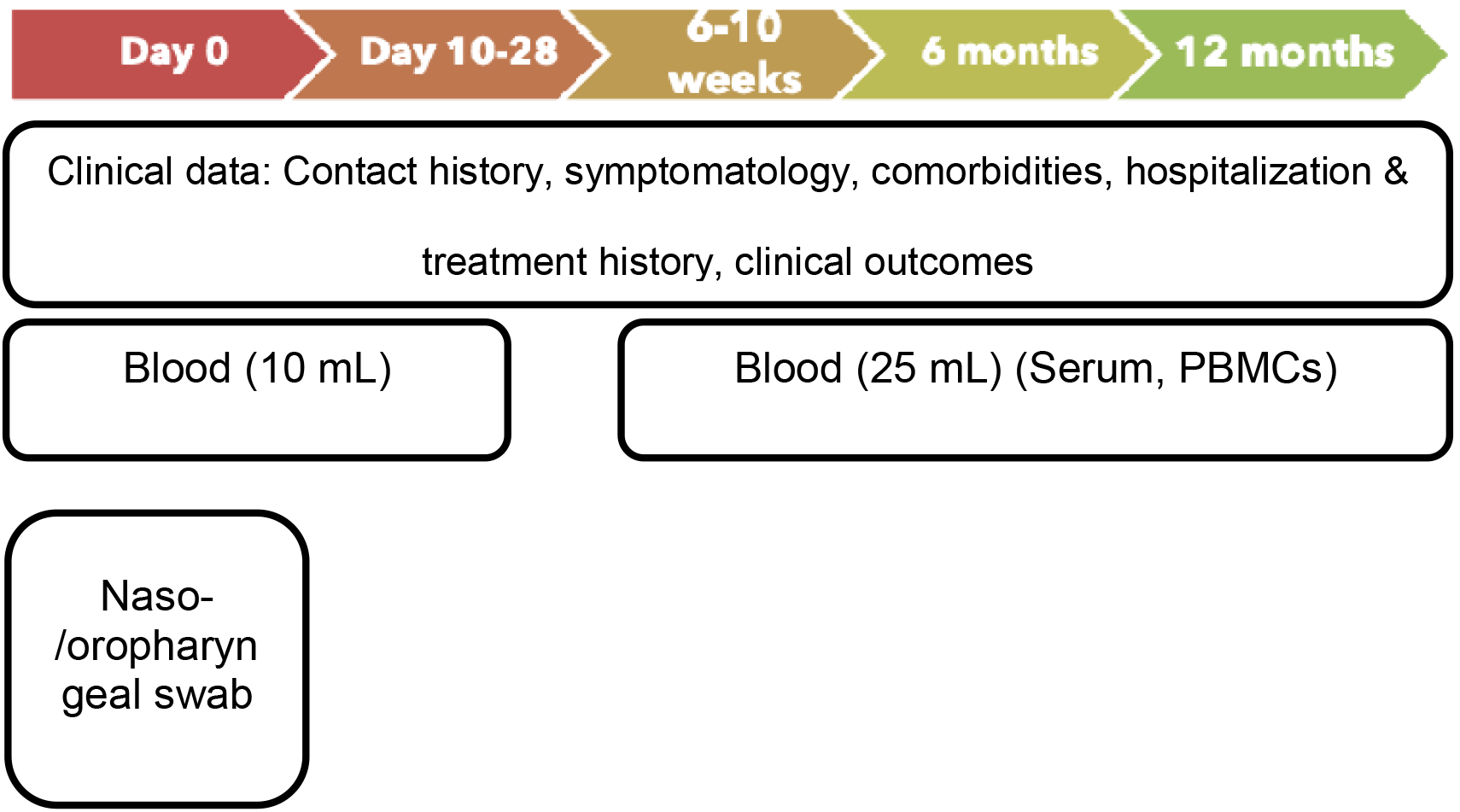
Study flow of the cohort. The follow up presented here is up to 6-10 weeks.

**Supplementary figure 2:**
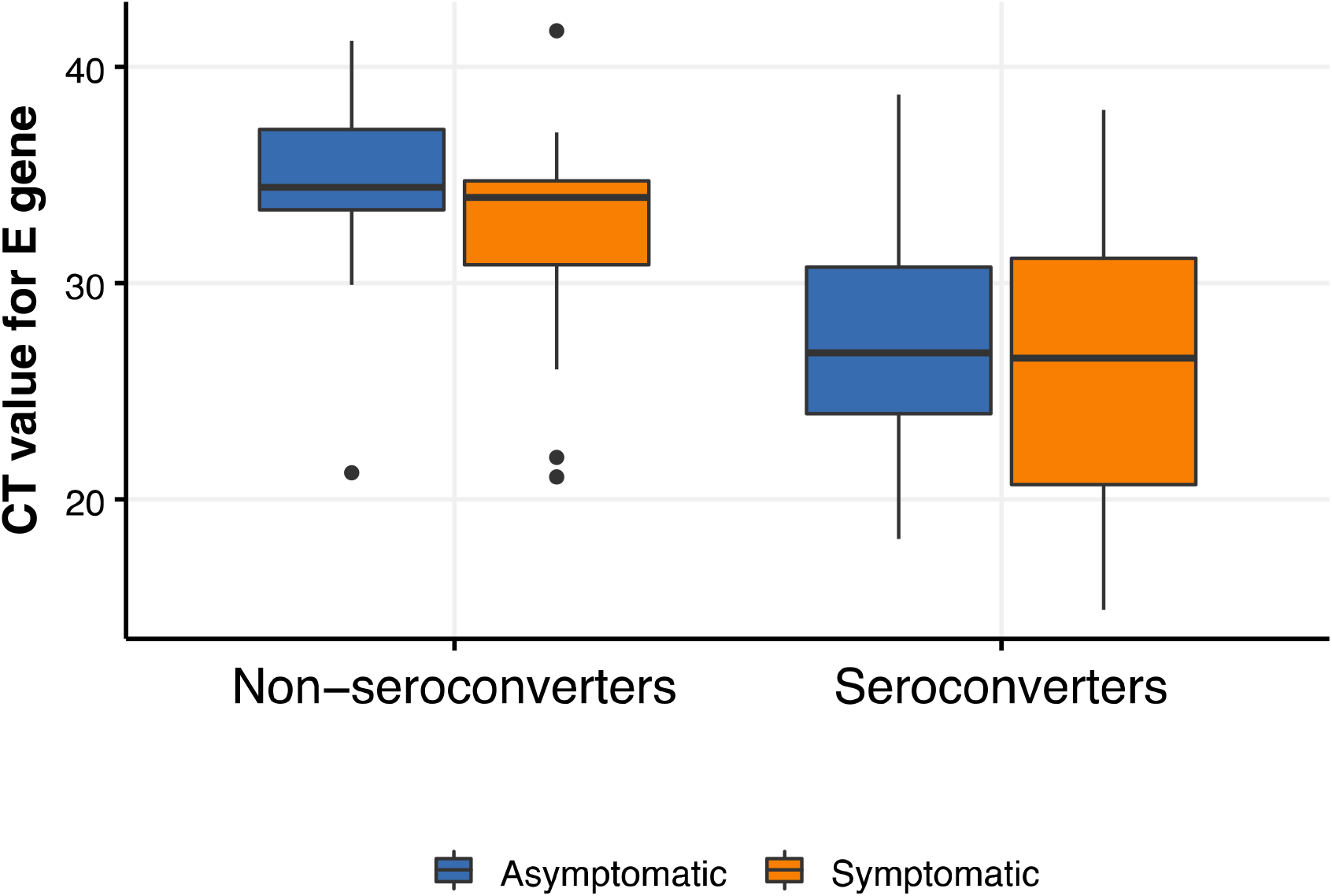
Comparison of RT-PCR threshold values (CT) between asymptomatic and symptomatic participants among seroconverters and non-seroconverters (N=169) False: Non seroconverters; True: Seroconverters Statistical test applied between asymptomatic and symptomatic groups: Mann Whitney test; among seroconverters: p-value 0.48 and among non seroconverters: p-value 0.16

**Table – S1.** Patterns of seropositivity across the three time-windows of illness.

| **Within 7 days**<br>**of onset of**<br>**illness\*** | **10-28 days** | **6-10 weeks** | **Anti-RBD**<br>**assay [n (%)]**<br>**N=746** | **Anti-NC assay**<br>**[n (%)]**<br>**N=673** |
| --- | --- | --- | --- | --- |
| Negative | Negative | Negative | 99 (13.3) | 120 (17.8%) |
| Negative | Negative | Positive | 21 (2.8) | 39 (5.8%) |
| Negative | Positive | Negative | 7 (1.0) | 12 (1.8%) |
| Negative | Positive | Positive | 305 (40.9) | 269 (40.0%) |
| Positive | Negative | Negative | 2 (0.3) | 1 |
| Positive | Negative | Positive | 1 (0.1) | 0 |
| Positive | Positive | Negative | 2 (0.3) | 12 (1.9%) |
| Positive | Positive | Positive | 309 (41.4) | 220 (33.7%) |

\*From the onset of symptoms in case of symptomatic or date of testing for asymptomatic participants
Anti-NC and anti-RBD IgG denote anti-nucleocapsid and anti-RBD Immunoglobulin-G respectively.

